# Machine learning for medication error detection: a scoping review protocol

**DOI:** 10.1101/2025.08.06.25333103

**Authors:** Félicien Hêche, Anthony Yazdani, Sohrab Ferdowsi, Ryme Kabak, Gang Mu, Douglas Teodoro

## Abstract

**Background:** Medication errors pose a significant threat to public health. Despite efforts by health agencies and the implementation of various interventions, such as staff training, medication reconciliation, and automation, the persistence of these incidents highlights the need for more effective, scalable solutions. In recent years, machine learning (ML) has emerged as a promising approach in healthcare, offering potential to detect and predict medication errors, through data-driven insights.

**Objective:** This scoping review aims to systematically map the existing literature on ML-based approaches to predict or detect medication errors across all stages of the medication use process. The review seeks to identify the range of ML applications in this domain, characterize methodological trends, and highlight current knowledge gaps. The findings will provide a structured and accessible overview for both clinicians and researchers, supporting the development of safer, more data-informed medication practices.

**Method and analysis:** The review will be conducted in accordance with the Preferred Reporting Items for Systematic Reviews and Meta-Analyses extension for Scoping Reviews (PRISMA-ScR) guideline. Structured searches will be performed in PubMed, Embase, and Web of Science. Predefined inclusion and exclusion criteria will be used to identify eligible studies. Key information – including ML model, data sources and type, evaluation methods, and clinical context – will be extracted and analyzed using descriptive statistics, visualizations, thematic analysis, and narrative synthesis.

**Study registration:** This protocol has been registered on the Open Science Framework (https://doi.org/10.17605/OSF.IO/38SFY).

## 1 Introduction

Medication errors—defined as failures in the treatment process that lead to, or have the potential to lead to, harm to the patient [1]—constitute a significant threat to public health worldwide. In England alone, more than 237 million medication errors are estimated to occur annually [2]. A study conducted in two adult outpatient cancer centers reported that these incidents affected 23% of prescribed drugs [3]. Additionally, approximately 3% of patients experience preventable harm related to medication use [4].

Medication errors can occur at any stage of the medication use process, including administration [5, 6], prescribing [7], or dispensing [8]. These errors arise not only in routine healthcare settings but also in clinical research, where complex protocols and investigational treatments may introduce additional risk factors [9, 10]. These incidents are inherently multifactorial, often resulting from interrelated factors such as fatigue [11, 12] or work overload [13]. Their consequences can range from minor inconveniences to serious adverse drug events [14, 15], including fatal outcomes [16, 17, 18]. Additionally, medication errors contribute to increased healthcare costs [2, 19] and prolonged hospital stays [20].

In response to this critical issue, several health agencies have launched campaigns to promote safer medication practices. The World Health Organization (WHO) introduced the Medication Without Harm initiative [21] and recently issued a policy brief to support its ongoing execution [22]. In England, the National Health Service (NHS) has identified medication safety as a key aspect of its Patient Safety Strategy [23], noting that drug-related incidents account for approximately 10% of all patient safety reports [24]. In the United States, the Food and Drug Administration (FDA) has long acknowledged the need to address medication errors, which led to the establishment of the Division of Medication Error Prevention and Analysis (DMEPA) in 1999 [25, 26]. This division is specifically dedicated to identifying, analyzing, and preventing medication errors in both premarket and postmarket stages of the drug development and regulatory process.

Various strategies have been explored to mitigate these preventable events [27, 28], including medication reconciliation [29, 30], staff training [31], the adoption of automated drug administration systems [32], or the development of packaging and labeling guidelines to reduce the risk of confusion between look-alike and sound-alike products [33]. However, these interventions face some drawbacks. For example, in clinical trials, investigational drugs may not have finalized commercial packaging or labeling, rendering design-focused strategies inapplicable. Automated medication administration systems require staff training and might cause technical issues [34]. Medication reconciliation entails extensive use of resources, organizational change, and interprofessional collaboration [35], limiting its implementation [36, 37]. Staff training can reduce the risk of medication errors, but financial challenges and workload pressures might limit its implementation [38].

Over recent years, machine learning (ML) methods have achieved notable success across a variety of domains, including computer vision [39, 40], natural language processing [41, 42, 43], and finance [44, 45]. In the healthcare sector, ML has demonstrated promising capabilities in applications such as medical image analysis [46, 47], early disease detection [48, 49], treatment recommendation [50, 51], and clinical trial risk assessment [52, 53, 54]. ML methods are particularly well-suited for learning from complex, and potentially multimodal datasets [55, 56]. As such, they hold strong promise for uncovering intricate relationships among patient characteristics, treatments, and outcomes—patterns that are frequently difficult to capture using traditional rule-based or statistical approaches, especially when working with large-scale data [57, 58]. Moreover, when integrated into healthcare workflows, ML models can provide real-time decision support, thereby contributing to safer and more efficient medical practice [59, 60]. Collectively, these capabilities underscore the promise of ML in the detection of medication errors, a challenge that has already motivated a growing body of research [61, 62, 63, 64, 65].

Since medication errors remain a major concern in clinical practice and research, a comprehensive understanding of how ML has been applied to detect them would be valuable to both researchers and healthcare professionals. For the scientific community, a scoping review could help synthesize current approaches, identify methodological trends, and highlight gaps for future investigation. For clinicians, it would offer an overview of existing tools and emerging technologies that could enhance medication safety. Previous work has highlighted the promise of ML in improving patient safety [66], and a recent scoping review has examined the use of ML in optimizing medication alerts—primarily during the prescribing phase [67]. However, to the best of our knowledge, no review has specifically addressed how ML has been applied across the broader spectrum of medication errors.

This work presents a scoping review protocol designed to systematically map the existing research on ML-based approaches to mitigate medication errors. Following the Preferred Reporting Items for Systematic Reviews and Meta-Analyses extension for Scoping Reviews (PRISMA-ScR) guideline [68], we will perform structured searches across PubMed, Embase, and Web of Science, applying predefined inclusion and exclusion criteria to identify relevant studies. Key information—including model, data sources and type, evaluation method, and clinical context—will be extracted and analyzed using descriptive statistics, visualizations, thematic analysis, and narrative synthesis. By providing a comprehensive overview of the current literature, this review aims to support clinicians in identifying relevant tools and guide researchers in advancing the application of ML to improve patient safety.

## 2 Methods and analysis

In this section, we present the protocol that will be used to conduct our literature review. Specifically, this protocol consists of five key stages: (i) formulation of the research questions; (ii) identifying relevant literature; (iii) study selection; (iv) data charting; and (v) synthesis and analysis of the results.

### 2.1 Stage 1: formulation of the research questions

The aim of this review is to identify how ML methods have been applied to predict or detect medication errors across various medical settings. To achieve this objective, the following research questions have been developed:

1. What types of ML methods have been used to predict or detect medication errors?
2. In which medical context have these approaches been applied?
3. What kinds of data sources and modalities have been utilized in these applications?
4. What are the main challenges, limitation, evaluation strategies, and reported outcomes?

### 2.2 Stage 2: identifying relevant literature

After careful consideration, we selected PubMed, Embase, and Web of Science as the primary databases for this review. The search strategy has been constructed using the following general query format:

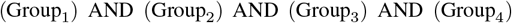

where each Group_*i*_ corresponds to a predefined set of keywords listed in Table 1. All terms were searched exclusively within the title and abstract fields.

**Table 1:** List of search terms included in each group.

| Group | Search Terms |
| --- | --- |
| Group 1 | drug error, medication error, contraindication, dosing error, dose error, prescription error, drug administration error, medication administration error, ordering error, route error, frequency error, strength error, formulation error, substitution error, preparation error, transcription error, dispensing error, monitoring error, omission error, commission error, labeling error, documentation error, storage error, wrong patient, wrong drug, wrong dose, wrong route, wrong time, medication safety, medication incident, drug safety |
| Group 2 | drug, medication |
| Group 3 | risk assessment, risk stratification, prediction, detection, classification, categorization |
| Group 4 | artificial intelligence, machine learning, deep learning, neural network, LSTM, long short-term memory, CNN, RNN, GRU, gated recurrent unit, autoencoder, multilayer perceptron, MLP, language model, decision tree, random forest, XGBoost, SVM, support vector machine, gradient boosting, LightGBM, adaptive boosting, AdaBoost, categorical boosting, CatBoost, supervised learning, unsupervised learning, reinforcement learning, ensemble learning, natural language processing, NLP |

Groups 1 and 2 were designed to capture literature related to medication errors. Specifically, Group 1 includes various types of errors, while Group 2 provides contextual keywords. Group 3 focuses on identifying studies involving risk assessment, prediction, detection or classification, and Group 4 targets literature that applies ML or artificial intelligence methodologies. The detailed PubMed keyword search is provided in Appendix A.

We considered articles published in English between January 1, 2015, and the date of the search (April 28, 2025). We also used available filters to automatically exclude literature reviews and non-peer-reviewed works, when possible. The initial search retrieved 306 records from PubMed, 252 from Embase, and 299 from Web of Science. Duplicate records were identified and removed from the initial set of 857 articles, using the reference management tool EndNote, resulting in a set of 581 studies.

### 2.3 Stage 3: study selection

In this stage of the review process, we will apply the eligibility criteria to the set of previously identified articles to select which studies will be included in the scoping review. This section outlines the inclusion and exclusion criteria and describes the procedure used to assess whether a specific article meets these requirements.

Studies are eligible for inclusion if they (i) develop, apply, or evaluate ML methods to predict or detect medication errors; (ii) are related to clinical treatment or research involving human subjects; (iii) are basic research articles published in peer-reviewed journals or conference proceedings, available in full text, and written in English; and (iv) have been published between 1 January 2015 and April 28, 2025.

We will exclude studies relying solely on statistical or rule-based approaches. In addition, articles that address adverse drug events without explicitly focusing on medication errors will be excluded. Works describing methods to prevent medication errors will be excluded unless they involve the prediction of such incidents. These inclusion and exclusion criteria are fully presented in Table 2.

**Table 2:** Review eligibility criteria based on concept, context, article type, and publication period.

|  | **Inclusion** | **Exclusion** |
| --- | --- | --- |
| **Concept** | Studies that develop, apply or evaluate ML methods to predict or detect medication errors. | Studies that do not use ML; studies relying solely on traditional statistical methods or rule-based systems; studies that propose preventive approaches that do not involve medication error predictions. |
| **Context** | Studies related to clinical treatment or research and involving human subjects. | Studies not related with clinical treatment or research; studies that do not involve human subjects. |
| **Types of articles** | Basic research articles published in peer-reviewed journals or conference proceedings; full-text available; written in English. | Reviews, editorials, opinion papers, abstracts, posters, dissertations, articles without full text, or not written in English. |
| **Time period** | Articles published between January 1, 2015 and April 28, 2025. | Articles published before January 1, 2015 or after April 28, 2025. |

The selection procedure will be carried out by two independent reviewers. First, titles and abstracts of the articles will be screened to assess their compliance with the eligibility criteria. Studies deemed potentially relevant will then undergo full-text screening. Those that meet the eligibility criteria upon full-text review will be included in the final set of studies for the scoping review. For studies where eligibility is unclear or ambiguous, inclusion decisions will be resolved through discussion and consensus among the two reviewers.

### 2.4 Stage 4: data charting

For each article included in the scoping review, we will extract key information to enable a comprehensive mapping of the existing literature. This includes metadata such as the title, authors, year of publication, and the journal or conference in which the article was published. We will also document the geographic location of the study and its main characteristics, including study design, objectives, and clinical context. In addition, we will extract details on the data used, including data sources, types, features, labels, and dataset size, as well as the specific ML models employed. Particular attention will be paid to the way ML methods are applied to predict or detect medication errors. Evaluation approaches (e.g., cross-validation, external validation) and performance metrics (e.g., accuracy, AUROC, precision, recall) will also be recorded. The complete set of extracted information is described in Table 3. This extraction framework has been developed with guidance from the CHecklist for critical Appraisal and data extraction for systematic Reviews of prediction Modelling Studies (CHARMS) [69].

**Table 3:** Description of the information extracted from each included study.

| Item | Information extracted |
| --- | --- |
| Title | Full title of the article. |
| Author(s) | Name of the first author. |
| Year | Year of publication. |
| Journal/Conference | Name of the journal or the conference where the article has been published. |
| Location | Geographic location where the study was conducted (if reported). |
| Study design | Type of study (e.g., retrospective study, prospective study, validation study, feasibility study). |
| Medication errors | Type of medication errors considered in the study (e.g., administration errors, prescription errors, dosing errors). |
| Study objective | Objective of the study (e.g., predicting administration errors, detecting prescribing errors). |
| Clinical context | Clinical setting and environment (e.g., hospital, outpatient clinic, pharmacy), therapeutic area or disease focus, target population characteristics (e.g., age group, risk profile), and clinical intervention or research. |
| Data sources and type | Origin of data (e.g., electronic health records, registry, clinical trial, claims), and data modality (e.g., structured data, medical imaging, free-text clinical notes, multi-modal). |
| Features | Description of the features and their construction process. |
| Labels | Description of the labels and their annotation process. |
| Dataset size | Number of data instances for training, validation, and testing, including any external datasets if applicable. |
| ML model(s) | ML technique(s) used (e.g., decision trees, random forests, LSTM, large language models). |
| Method | Description of how ML was used to predict or detect medication errors; specific task formulation (e.g., classification, regression, anomaly detection); integration into the clinical workflow if reported. |
| Evaluation method | Description of validation strategies (e.g., cross-validation, test set, external validation). |
| Performance | Reported performance metrics (e.g., accuracy, AUROC, precision, recall, F1-score). |
| Additional notes | Any other relevant information or observations not captured by the categories above. |

### 2.5 Stage 5: synthesis and analysis of the results

The results of the scoping review will be presented using multiple complementary methods to provide a comprehensive overview of research topics and methodological approaches employed in the included studies. The synthesis will begin with a descriptive summary, potentially incorporating numerical indicators such as the number of publications per year, types of data used, ML models applied, and medical contexts considered. To enhance interpretability, visual aids may be employed, including bar charts to illustrate distributions across key categories (e.g., ML algorithms, dataset sizes), and summary tables to concisely present extracted information related to use cases, model performance, and dataset characteristics.

Based on the findings of this exploratory phase, we will determine the most suitable analytical framework for synthesis. This may involve thematic analysis to identify, analyze, and interpret recurring themes across studies—such as application contexts, methodological challenges, and validation strategies.

Finally, a narrative synthesis will be provided to describe the scope and nature of ML approaches for predicting or detecting medication errors. This synthesis will highlight key research trends, methodological patterns, and gaps in the literature. All presentation methods may be adapted as needed to accommodate the heterogeneity and specificity of the included studies.

## 3 Discussion

Medication errors remain a critical challenge in modern healthcare, contributing to avoidable patient harm and increased healthcare costs [2, 19]. In the context of clinical research, these errors can also have serious consequences, including the potential to trigger clinical holds [70], thereby delaying drug development timelines and increasing associated costs. As ML methods gain traction in clinical settings, their potential to predict and detect these errors warrants systematic evaluation. This scoping review protocol outlines a structured and transparent approach to mapping the existing literature on ML applications for medication error mitigation, capturing the breadth of methodological strategies, clinical contexts, and implementation efforts reported to date.

A strength of this protocol is its alignment with the PRISMA-ScR framework, ensuring methodological rigor and reproducibility. The use of three major bibliographic databases (PubMed, Embase, and Web of Science), a clearly defined search strategy, and detailed eligibility criteria further support the comprehensive identification of relevant studies. In addition, the review’s data extraction plan, based on the CHARMS framework, has been carefully designed to address both clinical and technical dimensions of ML-based approaches, allowing for a nuanced and multi-faceted synthesis.

However, the protocol is subject to certain limitations. The review is restricted to articles published in English and available in full text, which may introduce language and publication biases. Furthermore, the heterogeneity of study designs and ML evaluation methods may hinder the comparability of the findings and the ability to identify the most effective approach.

Nevertheless, the anticipated outcomes of this review are valuable. By systematically mapping current applications of ML in medication error detection, the review will help highlight promising approaches and expose scientific gaps. This will offer researchers a foundation for future investigations and provide clinicians and healthcare decision-makers with an accessible overview of how ML can be leveraged to enhance patient safety across different stages of the medication use process.

## Data Availability

All data used in this study were openly available prior to its initiation. The data will be retrieved from the following public bibliographic databases: PubMed (https://pubmed.ncbi.nlm.nih.gov/), Embase (https://www.embase.com/), and Web of Science (https://www.webofscience.com/).

## Ethics and dissemination

This study involves a review of existing literature and does not involve human participants, personal data, or unpublished secondary data. As such, ethical approval was not required. All data analyzed were obtained from publicly available sources. Findings of the scoping review will be disseminated through professional networks, conference presentations and publication in a scientific journal.

## Contributions

All authors conceived the study protocol steps. FH developed the structure of the manuscript and led manuscript development. All authors (FH, AZ, SF, RK, GM, DT) reviewed several iterations of the manuscript and approved the final version.

## Funding statement

This work was supported by Innosuisse – the Swiss Innovation Agency – grant number 114.721 IP-ICT

## Competing interest statement

None declared.

## A Appendix

The following query has been used to perform our research in PubMed.

(

“drug error*”[Title/Abstract] OR “medication error*”[Title/Abstract] OR “contraindication*”[Title/ Abstract] OR “dosing error*”[Title/Abstract] OR “dose error*”[Title/Abstract] OR “prescri* error*”[Title/Abstract] OR “drug administrat* error*”[Title/Abstract] OR “medication administrat* error*”[Title/Abstract] OR “order* error*”[Title/Abstract] OR “route error*”[Title/Abstract] OR “frequency error*”[Title/Abstract] OR “strength error*”[Title/Abstract] OR “formulation error*”[Title/Abstract] OR “substitution error*”[Title/Abstract] OR “preparation error*”[Title/Abstract] OR “transcrib* error*”[Title/Abstract] OR “dispensing error*”[Title/Abstract] OR “monitoring error*”[Title/ Abstract] OR “omission error*”[Title/Abstract] OR “commission error*”[Title/Abstract] OR “labeling error*”[Title/Abstract] OR “documentation error*”[Title/Abstract] OR “storage error*”[Title/ Abstract] OR “wrong patient”[Title/Abstract] OR “wrong drug”[Title/Abstract] OR “wrong dose”[Title/ Abstract] OR “wrong route”[Title/Abstract] OR “wrong time”[Title/Abstract] OR “medication safety”[Title/Abstract] OR “medication incident*”[Title/Abstract] OR “drug safety”[Title/Abstract]

)

AND

(

“drug”[Title/Abstract] OR “drugs”[Title/Abstract] OR “medication*”[Title/Abstract]

)

AND

(

“risk assessment”[Title/Abstract] OR “risk stratification”[Title/Abstract] OR “prediction”[Title/Abstract] OR “detection”[Title/Abstract] OR “classif*”[Title/Abstract] OR “categor*”[Title/Abstract]

)

AND

(

“artificial intelligence”[Title/Abstract] OR “machine learning”[Title/Abstract] OR “deep learning”[Title/Abstract] OR “neural network*”[Title/Abstract] OR “LSTM”[Title/Abstract] OR “long short-term memory”[Title/Abstract] OR “CNN”[Title/Abstract] OR “RNN”[Title/Abstract] OR “GRU”[Title/Abstract] OR “gated recurrent unit*”[Title/Abstract] OR “autoencoder*”[Title/Abstract] OR “multilayer perceptron*”[Title/Abstract] OR “MLP”[Title/Abstract] OR “language model*”[Title/Abstract] OR “decision tree*”[Title/Abstract] OR “random forest*”[Title/Abstract] OR “XGBoost”[Title/Abstract] OR “SVM”[Title/Abstract] OR “support vector machine”[Title/Abstract] OR “gradient boosting”[Title/ Abstract] OR “LightGBM”[Title/Abstract] OR “adaptive boosting”[Title/Abstract] OR “AdaBoost”[Title/ Abstract] OR “categorical boosting”[Title/Abstract] OR “CatBoost”[Title/Abstract] OR “supervised learning”[Title/Abstract] OR “unsupervised learning”[Title/Abstract] OR “reinforcement learning”[Title/Abstract] OR “ensemble learning”[Title/Abstract] OR “natural language processing”[Title/ Abstract] OR “NLP”[Title/Abstract]

)

AND (english[Filter]) AND (2015:2025[pdat])

## Notes

### Competing Interest Statement

The authors have declared no competing interest.

### Clinical Protocols

https://doi.org/10.17605/OSF.IO/38SFY

### Author Declarations

All data used in this study were openly available prior to its initiation. The data will be retrieved from the following public bibliographic databases, PubMed (https://pubmed.ncbi.nlm.nih.gov/), Embase (https://www.embase.com/), Web of Science (https://www.webofscience.com/)

